# Performance of ChatGPT on free-response, clinical reasoning exams

**DOI:** 10.1101/2023.03.24.23287731

**Authors:** Eric Strong, Alicia DiGiammarino, Yingjie Weng, Preetha Basaviah, Poonam Hosamani, Andre Kumar, Andrew Nevins, John Kugler, Jason Hom, Jonathan H Chen

## Abstract

**Importance:** Studies show that ChatGPT, a general purpose large language model chatbot, could pass the multiple-choice US Medical Licensing Exams, but the model’s performance on open-ended clinical reasoning is unknown.

**Objective:** To determine if ChatGPT is capable of consistently meeting the passing threshold on free-response, case-based clinical reasoning assessments.

**Design:** Fourteen multi-part cases were selected from clinical reasoning exams administered to pre-clerkship medical students between 2019 and 2022. For each case, the questions were run through ChatGPT twice and responses were recorded. Two clinician educators independently graded each run according to a standardized grading rubric. To further assess the degree of variation in ChatGPT’s performance, we repeated the analysis on a single high-complexity case 20 times.

**Setting:** A single US medical school

**Participants:** ChatGPT

**Main Outcomes and Measures:** Passing rate of ChatGPT’s scored responses and the range in model performance across multiple run throughs of a single case.

**Results:** 12 out of the 28 ChatGPT exam responses achieved a passing score (43%) with a mean score of 69% (95% CI: 65% to 73%) compared to the established passing threshold of 70%. When given the same case 20 separate times, ChatGPT’s performance on that case varied with scores ranging from 56% to 81%.

**Conclusions and Relevance:** ChatGPT’s ability to achieve a passing performance in nearly half of the cases analyzed demonstrates the need to revise clinical reasoning assessments and incorporate artificial intelligence (AI)-related topics into medical curricula and practice.

## Introduction

ChatGPT is a chatbot interface for the GPT-3 large language model artificial intelligence (AI) system that generates human-like text in response to user text input^1^. ChatGPT is already capable of approaching or exceeding the passing threshold for multiple-choice questions that simulate the United States Medical License Exams (USMLE)^2,3^. We examine herein how well the model responds to free-response, case-based questions with more general implications for the application, instruction, and assessment of clinical reasoning skills.

## Methods

We selected 14 clinical cases used for clinical reasoning final exams for first and second year medical students at our academic medical center from 2019 to 2022. Each exam consists of two cases on which students need to achieve a cumulative score of 70% to pass. Each case consists of a vignette providing data in discrete, sequential passages, separated by 2-7 free-response questions that assess a wide variety of specific clinical reasoning skills (Figure 1, Table 1). Each case was run through ChatGPT twice, and two faculty independently graded each response according to the rubrics used on the original exams. The mean from the two graders was the final score for a given run. Consistent with the student exams, the passing threshold was predefined as ≥ 70% for a given case. To assess variation in ChatGPT’s performance across multiple runs, we selected one high-complexity case to repeatedly run through the process 20 times.

**Figure 1:**
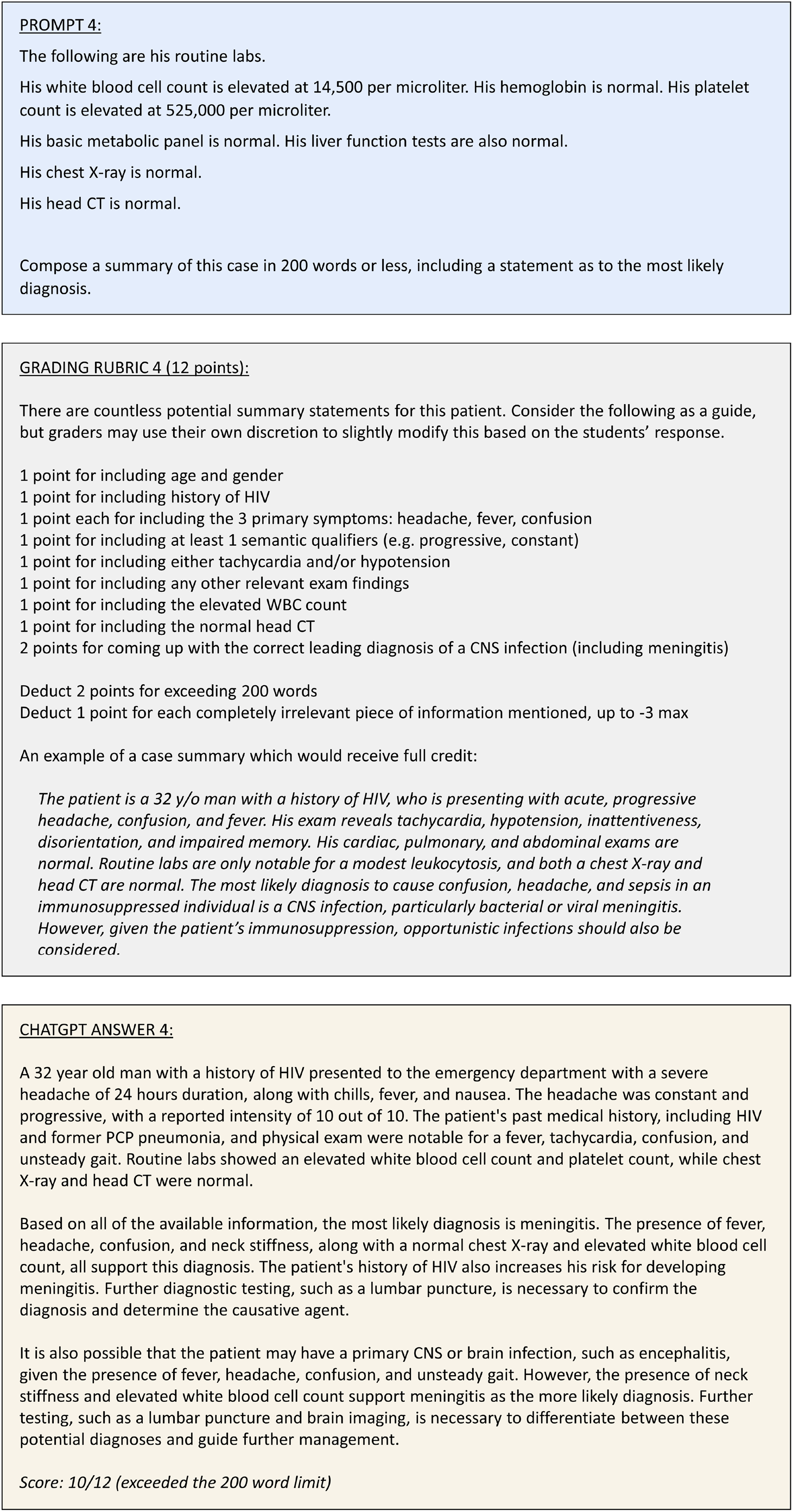
Representative example of a prompt from a clinical reasoning case. An excerpt from a representative example of the format of the clinical cases given to ChatGPT. There is prompt consisting of the text provided to ChatGPT verbatim. the standardized grading rubric, ChatGPT’s actual response, and the score assigned to that response by a grader.

**Table 1:** Summary of cases, clinical skills assessed, and scores for ChatGPT’s responses

| Case Description | Assessed Clinical Skills |  |  |  |  |  |  | Word Count | ChatGPT Score Run 1<sup>d</sup> | ChatGPT Score Run 2<sup>d</sup> |
| --- | --- | --- | --- | --- | --- | --- | --- | --- | --- | --- |
|  | Diagnostic schema<sup>a</sup> | Differential Diagnosis | Illness Scripts<sup>b</sup> | Test Interpretation | Assessment | Problem List | Other<sup>c</sup> |  |  |  |
| Chronic fatigue & anemia | X | X |  |  |  | X | X | 583 | 60.7% | 65.0% |
| Acute abdominal pain & diarrhea | X | X |  | X |  | X |  | 798 | 67.6% | 77.8% |
| Acute confusion & hypertension | X | X | X | X | X | X |  | 1013 | 69.5% | 61.3% |
| Chronic diarrhea & amenorrhea |  | X |  | X | X |  |  | 1109 | 62.7% | 68.5% |
| Subacute fever & abdom. pain | X | X |  | X |  | X | X | 919 | 62.8% | 73.0% |
| Chronic dyspnea |  | X |  |  |  | X |  | 831 | 60.7% | 60.1% |
| Acute chest pain |  |  |  |  |  |  | X | 747 | 81.3% | 83.3% |
| Acute RUQ pain |  | X |  | X | X |  | X | 902 | 58.8% | 64.2% |
| Acute lightheadedness |  | X |  | X | X | X |  | 885 | 40% | 43.0% |
| Acute abdominal pain & fever |  | X |  |  | X |  | X | 1071 | 81.1% | 77.8% |
| Chronic fatigue |  | X |  | X | X | X |  | 917 | 73.3% | 69.4% |
| Subacute confusion | X | X | X | X | X | X |  | 953 | 68.6% | 74.6% |
| Acute abdominal pain & nausea | X | X |  | X |  | X | X | 972 | 82.4% | 79.9% |
| Subacute dyspnea |  | X | X | X |  | X |  | 1093 | 77.0% | 84.7% |

a A diagnostic schema is defined as a thorough collection of etiologies for a specific symptom which is organized into categories based on organ system or physiologic process.
b An illness script is defined as a mental summary of features of a specific disease, organized into categories such as epidemiology, historical features, exam findings, and relevant test abnormalities.
c Other assessed clinical skills include: diagnostic test selection, identification of cognitive biases, discussion of relevant literature search strategies, and interpretation of the significance of physical exam findings.
d Scores listed are the mean score from two independent faculty graders using the same grading rubric.

## Results

ChatGPT met or exceeded the predefined passing threshold of 70% on 12 out of the 28 (43%) runs (Table 1), with a mean score of 69% (95% CI: 65% to 73%).

For the high-complexity case, the mean score was 68% (95% CI: 65% to 72%), and ChatGPT scored over 70% on 7 out of the 20 (35%) runs. For this case, ChatGPT’s performance varied between questions depending on the clinical reasoning task assessed. It performed best on the question that required the creation of relevant illness scripts, scoring 80% (95% CI: 74% to 86%); and performed worst on the question that required the creation of a relevant diagnostic schema, scoring 62% (95% CI: 55% to 68%).

## Discussion

Previous studies have demonstrated the ability of generative AI to perform well on multiple-choice examinations. Our study demonstrates ChatGPT’s potential to also reach the passing threshold on open-ended clinical reasoning exams – a significant AI milestone bringing challenges and opportunities.

An immediate problem for medical training concerns the reliability of formal assessments of students. A passing grade on a clinical reasoning final exam is an important benchmark in medical training – one that signals a student is sufficiently prepared for clerkships with real patients. The use of a chatbot to obtain a borderline passing grade for a student who otherwise would not have passed reduces the ability to identify trainees in need of remediation or other support to ensure subsequent success in providing safe and reliable patient care. The optimal approach will be to redesign assessments in order to retain the ability to identify struggling students despite the use of a chatbot. Such revisions will take time, so switching to in-person, closed-book exams may be a necessary, stop-gap measure. However, closed-book exams do not test the ability to integrate information from a variety of sources, and goes against the trend in medical assessments, such as the American Board of Internal Medicine allowing the use of UpToDate during recertification examinations^4^. Current and future physicians need a basic understanding of this technology, including the advantages and disadvantages, just as they had to learn the effective use of internet search and electronic medical records.

A limitation of this study is that ChatGPT’s responses can be sensitive to relatively minor rewording of prompts. For example, it demonstrated a different understanding of several specific clinical reasoning terms (e.g. illness script, problem list) as compared to those we use with our students. This required rewording of some questions to include an explanation of the relevant term. The bot may well have performed even better with additional trial-and-error in question phrasing. Another limitation is that the specific format of our clinical reasoning assessments may not resemble those used at all medical schools; however, the core skills tested are common throughout medical training.

Given the demonstrated abilities of general purpose chatbot AI systems, a broader incorporation of AI-related topics into medical training and practice has now become necessary. This rapidly advancing technology is likely to reshape the nature of education, assessment, and application of medical knowledge in practice.

## Data Availability

All data produced in the present study are available upon reasonable request to the authors

## Author Contributions

Dr. Strong had full access to all of the data in the study and takes responsibility for the integrity of the data and the accuracy of the data analysis.

Concept and design: Strong, DiGiammarino, Hom, Chen

Acquisition of data: Strong, DiGiammarino, Basaviah, Hosamani, Kumar, Hom

Authorship of cases: Nevins, Kugler, Strong

Statistical analysis: Weng

Drafting of the manuscript: DiGiammarino, Strong, Weng

Critical revision of the manuscript for important intellectual content: Hom, Chen

Administrative and organizational support: DiGiammarino

Supervision: Hom, Chen

Dr. Strong and Ms. DiGiammarino contributed equally as co-first authors.

Dr. Hom and Dr. Chen contributed equally as co-last authors.

## Acknowledgements

We would like to thank Madika Bryant, MA (Office of Medical Education, Stanford University School of Medicine) for her assistance with organizing the clinical cases and with recommendations for prior revisions to case grading rubrics.

## References

1. OpenAI. ChatGPT: Optimizing Language Models for Dialogue. OpenAI. Published November 30, 2022. https://openai.com/blog/chatgpt/

2. Gilson A, Safranek C, Huang T, et al. How Does ChatGPT Perform on the Medical Licensing Exams? The Implications of Large Language Models for Medical Education and Knowledge Assessment. Published online December 26, 2022. doi:10.1101/2022.12.23.22283901

3. Kung TH, Cheatham M, Medenilla A, et al. Performance of ChatGPT on USMLE: Potential for AI-Assisted Medical Education Using Large Language Models. Published online December 20, 2022. doi:10.1101/2022.12.19.22283643

4. Doctors maintaining ABIM Board Certification will soon be able to access an electronic resource they use in practice during periodic knowledge assessments. American Board of Internal Medicine. September 27, 2017. https://www.abim.org/media-center/press-releases/abim-open-book-assessments-will-feature-access-to-uptodate.aspx. Accessed February 2, 2023.

